# NATIONAL SCALE REAL-TIME SURVEILLANCE OF SARS-COV-2 VARIANTS DYNAMICS BY WASTEWATER MONITORING IN ISRAEL

**DOI:** 10.1101/2021.12.26.21268420

**Authors:** Itay Bar-Or, Victoria Indenbaum, Merav Weil, Michal Elul, Nofar Levi, Irina Aguvaev, Zvi Cohen, Virginia Levy, Roberto Azar, Batya Mannasse, Rachel Shirazi, Efrat Bochris, Neta S. Zuckerman, Alin Sela Brown, Danit Sofer, Orna Mor, Ella Mendelson, Oran Erster

## Abstract

In this report, we describe a national-scale monitoring of the SARS-COV-2 (SC-2) variant dynamics in Israel, using multiple-time sampling of twelve wastewater treatment plants. We used a combination of inclusive and selective quantitative PCR assays that specifically identify variants A19 or B.1.1.7 and tested each sample for the presence and relative viral RNA load of each variant. We show that between December-2020 and March-2021, a complete shift in the SC-2 variant circulation was observed, where the B.1.1.7 replaced the A19 in all examined test points. We further show that the normalized viral load (NVL) values and the average new cases per week reached a peak in January 2021, and then decreased gradually in almost all test points, in parallel with the progression of the national vaccination campaign, during February-March 2021. This study demonstrates the importance of monitoring SC-2 variant dynamics on a national scale through wastewater sampling. It also provides a proof-of-concept methodology for continuous surveillance by using a combination of inclusive and selective PCR tests, which is far more amendable for high throughput monitoring compared with sequencing. This approach may be useful for real-time dynamics surveillance of current and future variants, such as the Omicron (BA.1) variant.

**Synopsis:** This study describes the continuous monitoring of the SARS CoV-2 variant B.1.1.7 circulation in wastewater in Israel using a positive/negative quantitative PCR assay.

## 1. INTRODUCTION

Since its emergence in December 2019, SARS-COV-2 (SC-2) spread worldwide, causing the COVID19 pandemic, with unprecedented impact on human lives, economy, social aspects and public health throughout the world^1^. While SC-2 infection may cause a range of respiratory, gastrointestinal and systemic symptoms, the majority of individuals show little to no symptoms ^2–7^Clinical testing of individuals for SC-2 is the primary surveillance method for obtaining information for implementation of public health strategic interventions, such as quarantine, to mitigate the spread of the virus. The current gold standard for clinical testing is reverse transcriptase quantitative polymerase chain reaction (RT-qPCR)^3^, which detects the viral RNA. Wastewater samples have been utilized to identify several pathogenic human viruses and, consequently, it has gained attention for assessing population-level trends of SC-2 infections.

Detection of SC-2 in wastewater was reported in numerous studies, including North America^8–11^, Europe^12–16^, Asia^17,18^ and Oceania^19^, with feasibility addressed in the review by Mousazadeh et al.^5^. These studies describe a range of sample concentration and viral RNA recovery approaches, followed by RT-qPCR to determine the viral load. These proof-of-concept studies indicate that wastewater monitoring can be used as an early detection tool for health autorities^20^.

The emergence of SC-2 variants that have the potential for increased transmissibility and/or immune response evasion raised an urgent need for targeted surveillance of circulating lineages. SC-2 variant B.1.1.7 (https://nextstrain.org/sars-cov-2/, Alpha variant) was first identified in the UK during December 2020, where it became the dominant SC-2 within four weeks^6^. The genome of variant B.1.1.7 contains 23 mutations, some of which are associated with increased transmissibility, which can lead to higher viral loads in infected patients, and subsequent increased mortality^21^. The dynamic shifts in SC-2 variant dominance require enhanced surveillance of emerging lineages such as B.1.1.7., to facilitate appropriate response by public health authorities. The ultimate tool for identification of new variants is the Whole genome sequencing (WGS) approach^22,23^. However, its use is time-consuming, expensive, requires highly skilled personnel, and cannot be readily scaled up to accommodate a large number of samples. Methods based on RT-qPCR, which include a “drop-out” signal, available in commercial kits (such as the Thermo Scientific TaqPath COVID-19 diagnostic kit) or the method published in a recent study^24^ are cost-effective, rapid and can be readily scaled up in any molecular biology laboratory. This approach has a major limitation, which stems from the issues associated with absence of signal. Such a “drop out” can result from low sensitivity or inhibition, and may not necessarily reflect the presence of the target mutation. Furthermore, in the case of the Thermo Scientific kit, the 69-70 mutation, which is the cause of the “drop out” effect, is not unique to variant B.1.1.7., and cannot be considered as a reliable indication of this variant. Other commercial kits that target specific SC-2 mutations are more specific, but require complicated interpretation, and in some cases, multiple tests per sample, to determine the sample identity (https://www.kogene.co.kr/eng/, https://www.seegene.com/, en.vircell.com).

The third morbidity wave of COVID19 in Israel started in mid-November 2020. In mid-December 2020, the first cases of SC-2 alpha variant were detected in Israel^25^. As a result, there was a shift in the SC-2 variant dominance, from A19/A20 (“WT” Wild type, Wuhan strain) to B.1.1.7 (alpha) variant in Israel. The national vaccination campaign (using the BNT162b2 Pfizer vaccine) started at the end of December 2020, and by the end of March 2021, 56% and 51% of the total population were vaccinated with the first and the second doses, respectively (https://datadashboard.health.gov.il/COVID-19/general).

In a previous report, we described the development of a PCR assay that identifies a mutation unique to variant B.1.1.7., and showed that it can be used as a reliable marker for its presence of the unknown samples^26^. Here, we combined this assay with a complementing test, so that the relative load of either B.1.1.7. or non-B.1.1.7. RNA in an examined sample can be evaluated. This study demonstrates the use of a rapid test to monitor the SC-2 variant dynamics on a national scale in real time, by continuous sampling of wastewater.

## 2. MATERIALS AND METHODS

### 2.1. Wastewater sampling and processing

The wastewater sampling sites (n=13) selected for this study were collected from various regions of Israel covering more than 55% of Israel population. These samples were collected from wastewater treatment plants (WWTP) between December 2020 and March 2021 (n=117 samples). In each sampling cycle, 250 ml of untreated wastewater was collected every 30 minutes for 24 hours, by external automatic composite samplers. Samples were transported to the laboratory under cooled conditions (4°C). The sample material was concentrated as previously described^19^, with some minor changes. A homogeneous fraction of the wastewater sample (25 ml) was centrifuged (4696 g) for 5 min at 4°C, and 20 ml of the supernatant was collected into 50 ml tube containing 0.26M of MgCl_2_. The tubes were gently stirred for 5 min. Then the sample was passed through 0.45-μm pore-size, 47-mm diameter electronegative MCE membranes (Merck Millipore Ltd) by peristaltic pump. The membrane was transferred into a 50 ml tube containing 3 ml external lysis buffer (NucliSENS easyMAG). The tube with the membrane was gently stirred for 30 min for virus inactivation and removal.

### 2.2. Viral RNA extraction

Total nucleic acid (NA) were extracted using the NucliSENS easyMAG system (Biomerieux, Marcyl’Etoile, France) following the manufacturer’s instructions. Briefly, 3 ml of lysis buffer containing the inactivated virus concentrated sewage were extracted using the easyMAG machine. Extracted NA were eluted using 55μL elution buffer and stored at -70°C.

### 2.3. Primers and probes

Primers and probe for the OC43 spike control reaction were from Dare et al.^27^. Primers and probe for the SC-2 Envelope (E) gene reaction were based on the sequences published by Corman et al.^28^ with the modifications detailed in **Table 1**. The primers for the SC-2 Nucleocapsid (N) gene reaction were based on the CDC nCov-N1 reaction (https://www.cdc.gov/coronavirus/2019-ncov/lab/rt-pcr-panel-primer-probes.html), with the modification detailed in **Table 1**. The B.1.1.7 specific reaction was based on the CDC nCoV-N1, with modifications described previously^26^. The WT-specific N1 reaction was termed N_WT_, and the alpha-specific reaction was termed N_B.1.1.7_ hereafter.

**Table 1.**
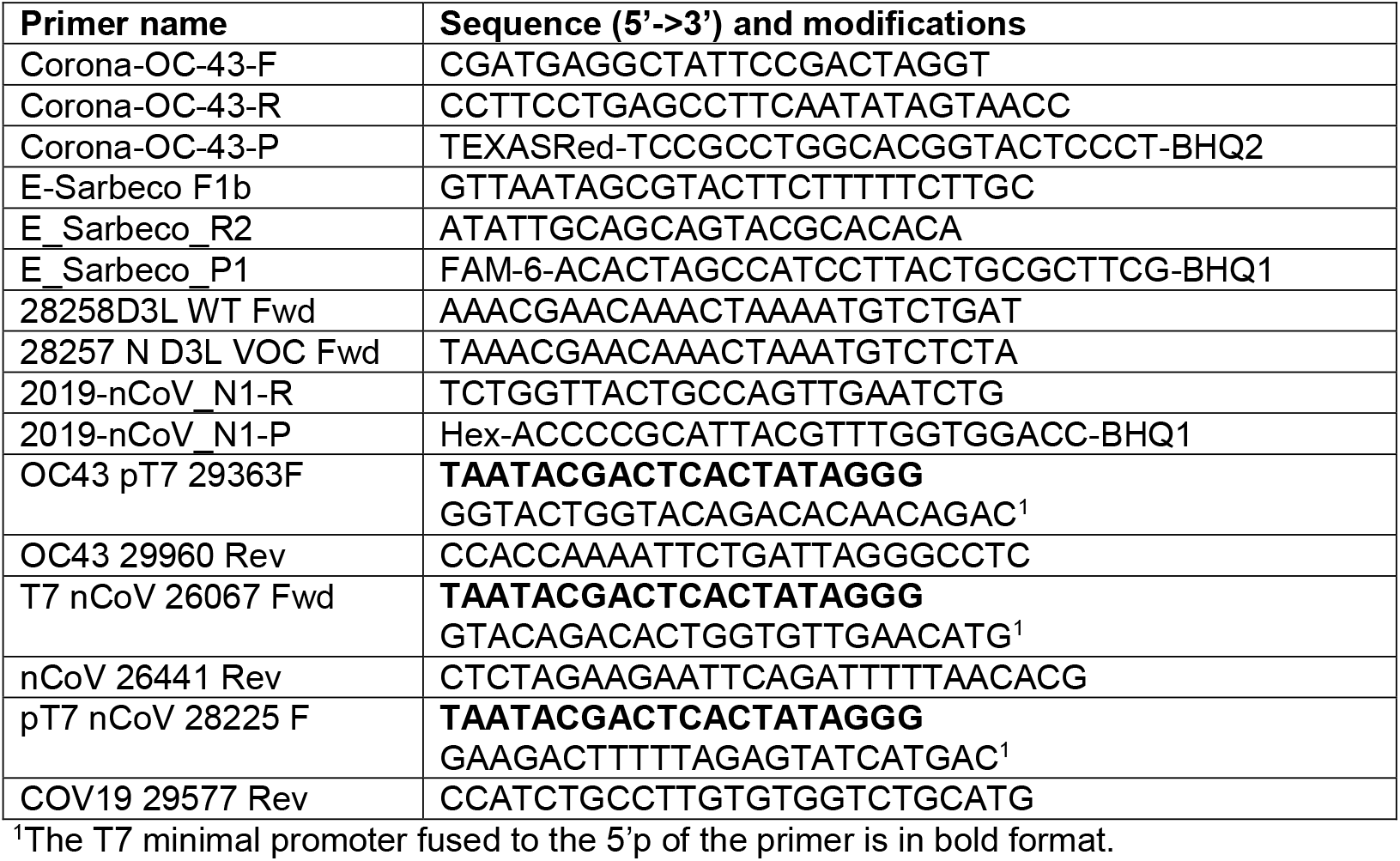
Primers and probes used in this study.

### 2.4. RT-qPCR

RT-qPCR mix was prepared using the Meridian (formerly Bioline) SensiFast one-step mix (https://www.bioline.com/sensifast-probe-no-rox-one-step-kit.html), with the addition of PCR-grade Bovine Serum Albumine (BSA) at a final concentration of 300nM. The reaction was run using Bio-Rad CFX96 instrument (https://www.bio-rad.com/en-us/product/cfx96-touch-real-time-pcr-detection-system?ID=LJB1YU15). The cycling conditions were as follows: 1 45.0°C for 15 minutes, 95.0°C for 2:20 minutes, 45X [95.0°C for 5 seconds, 60.0°C for 42 seconds]. Fluorescence was read at 60°C on each cycle. Analysis of the results were performed using the Bio-Rad CFX Maestro software (https://www.bio-rad.com/en-us/product/cfx96-touch-deep-well-real-time-pcr-detection-system?ID=LZJTUJ15).

### 2.5. Design and generation of In vitro transcribed standard RNA templates

*In-vitro* transcribed RNA molecules were used as control templates to maintain uniformity and to establish the reaction analytical Limit of Detection (LOD). Generation of RNA templates was performed using the Megascript kit (Thermo Fisher, https://www.thermofisher.com/order/catalog/product/AMB13345#/AMB13345), which allows *In-vitro* synthesis of RNA from DNA template using the T7 phage DNA dependent RNA polymerase. Regions flanking the target sequences of each reaction were amplified using specific primers fused to the T7 phage minimal promoter sequence, and were then used as template for the RNA synthesis. The resulting RNA templates were purified using the PSS magLEAD instrument (http://www.pss.co.jp/english/product/magtration/lead6-12gc.html). The RNA yield was determined using the NanoDrop spectrophotometer (https://www.thermofisher.com/i) and used to generate standard curves for each reaction. The analytical limit of detection (LOD) was determined using serial dilutions of the RNA targets, with the multiplex mix. This calibration was then used to calculate the number of copies detected in wastewater samples.

### 2.6. Calculation of the Normalized Viral Load value in wastewater samples

In order to compare the viral load as measured by qPCR, in different catchment points that have different flow rates and are draining regions with varying population size, we used a Normalized Viral Load (NVL) conversion that takes into account these variables^29^. Using standard curves generated with the *In vitro* transcribed standard RNA templates, qPCR Cq values of the examined samples were converted into viral RNA copies. These values were then used for the NVL calculation, according to the following equation:

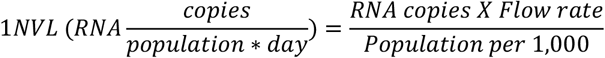

Where RNA copies per liter of wastewater is the converted from Cq value, Flow rate is the measured flow rate (liter per day) as measured in the specific catchment point, and Population per 1,000 is the size of the population in thousands, which reside in the region that the specific catchment point is collecting.

Calculation of the average NVL for each region was performed by determining the average NVL from each month (December, January, February and March) for each WWTP. Then, the average NVL values from all the points of each region were summarized for each month. NVL values based the copy number derived from the E gene reaction were calculated to determine the total NVL of SC-2 in the sample pool. The percentage of the A19/B19 RNA, and the B.1.1.7 RNA were determined by the NVL values of N_WT_ reaction and N_B.1.1.7_, respectively.

## 3. RESULTS

### 3.1. Development of the positive/negative B.1.1.7 variant reaction

The reaction that specifically detects the B.1.1.7 variant was described previously^26^. Briefly, a primer that binds specifically to the D3L substitution was designed and used. This mutation is almost exclusive to variant B.1.1.7 and is not currently associated with any other major SC-2 variant, as evident from the NextStrain global analysis (**Supplementary Figure S1A**). In order to perform a reciprocal reaction that detects only the WT lineage, a reciprocal primer that binds only the WT sequence was designed. The sequences and primer binding sites are illustrated in **Supplementary Figure S1B**. In order to monitor the on-going spreading of variant B.1.1.7 in Israel, the N_B.1.1.7_ assay was applied to wastewater samples, in parallel with its reciprocal N_WT_ reaction. For continuous monitoring of the variant circulation in wastewater, each sample was tested with a multiplex containing the E, N_WT_ and OC43 reactions, and again, with the E and N_B.1.1.7_ reactions. This enabled us to evaluate the overall SC-2 circulation in the examined wastewater treatment plants (WWTP), and at the same time, determine the percentage of WT versus B.1.1.7 variants in that WWTP.

### 3.2. Development of RT-qPCR assay for detection of SC-2 in environmental samples

The designed primers were required to detect the presence of SC-2, to distinguish between the WT and variant B.1.1.7 strains, and to confirm the integrity of the sample preparation. In order to meet these requirements, the following triplex qPCR assay was developed. An inclusive SC-2 reaction, targeting a conserved sequence within the SC-2 Envelop (E) gene was used as a control for the presence or absence of SC-2 RNA in the sample. A second, differential reaction, targeting the N gene B.1.1.7 variant-associated mutation D3L was used as the differential reaction, where the variant-specific primer was altered, depending on the reaction specificity. A third reaction, targeting the hCoV OC43, was used as an internal spiked control, to evaluate the pre-extraction procedure integrity.

In order to evaluate the specificity of the differential N_D3L_ reaction, 19A/19B and B.1.1.7 samples were examined, each using either the N_WT_ reaction or the N_B.1.1.7_ reaction. As shown in **Figure 1**, the 19A/19B sample was positive for both the E and N targets, when using the N_WT_ reaction, but only for the E target, when using the N_B.1.1.7_ reaction. The B.1.1.7 sample gave the reciprocal result, i.e. positive for the E target only with the N_WT_ reaction, and positive for both targets with the N_B.1.1.7_ reaction (**Figure 1**). Next, the components of the three reactions were combined together and their performance were tested using *In vitro* synthesized RNA targets. Serial dilutions of the RNA targets were spiked into wastewater extraction, to simulate an authentic sample background medium. The analytical limit of detection (LOD) derived from the dilutions testing was determined as follows: 35 copies per reaction for the E reaction, 12.9 copies for the N_B.1.1.7_ reaction, 13.5 copies for the N_WT_ reaction and 28 copies for the OC43 reaction (**Figure 2**).

**Figure 1.**
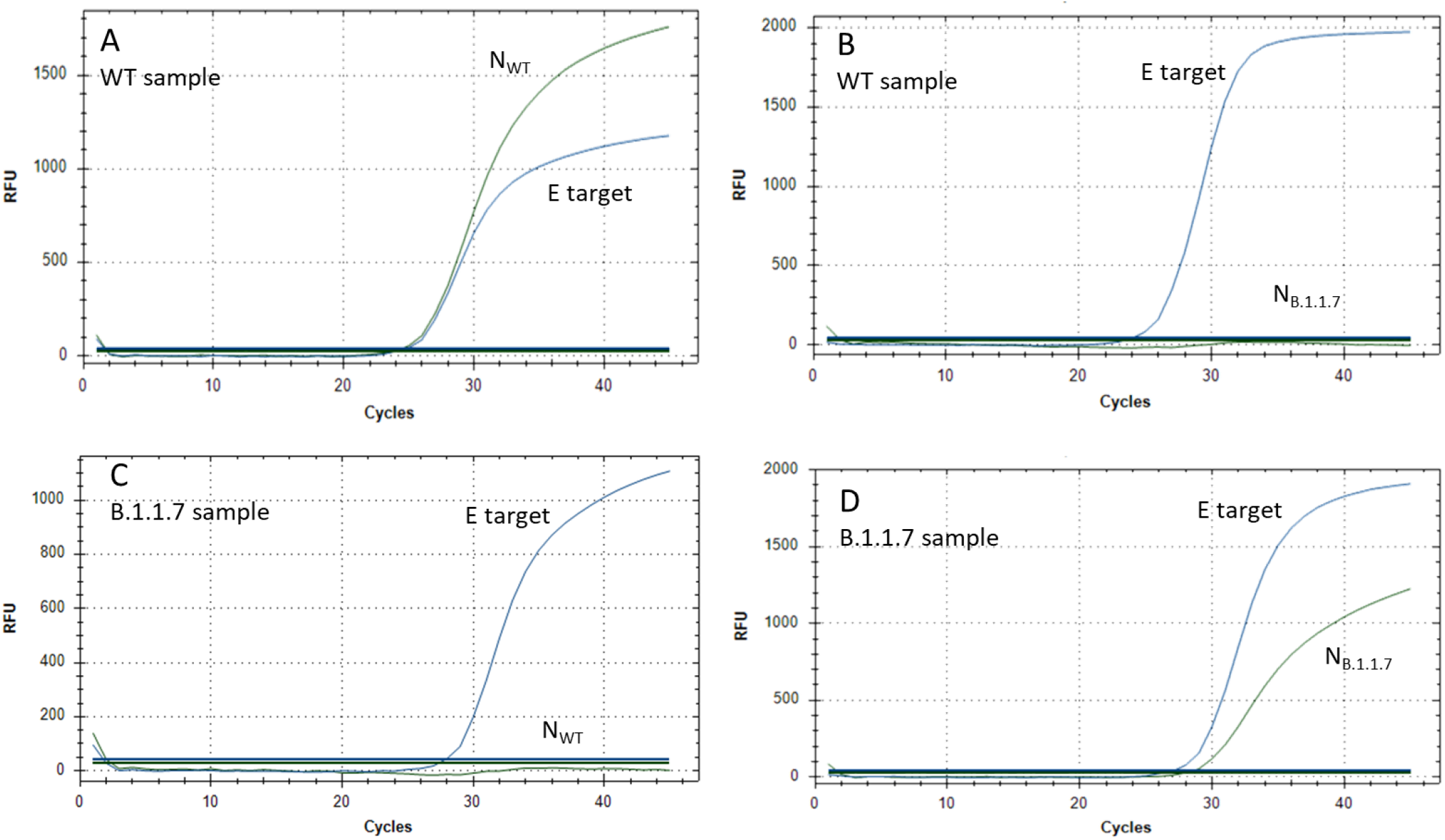
Amplification curves of the duplex SC-2 reactions. Each of the multiplex reactions was used with either A19/B19 (“WT”) sample or B.1.1.7 (alpha) sample. (A) WT reaction with WT sample. (B) Alpha reaction with WT sample. (C) WT reaction with alpha sample. (D) Alpha reaction with alpha sample. WT reaction: Multiplex detecting the E and N_WT_ targets. Alpha reaction: Multiplex detecting the E and N_B.1.1.7_ targets.

**Figure 2.**
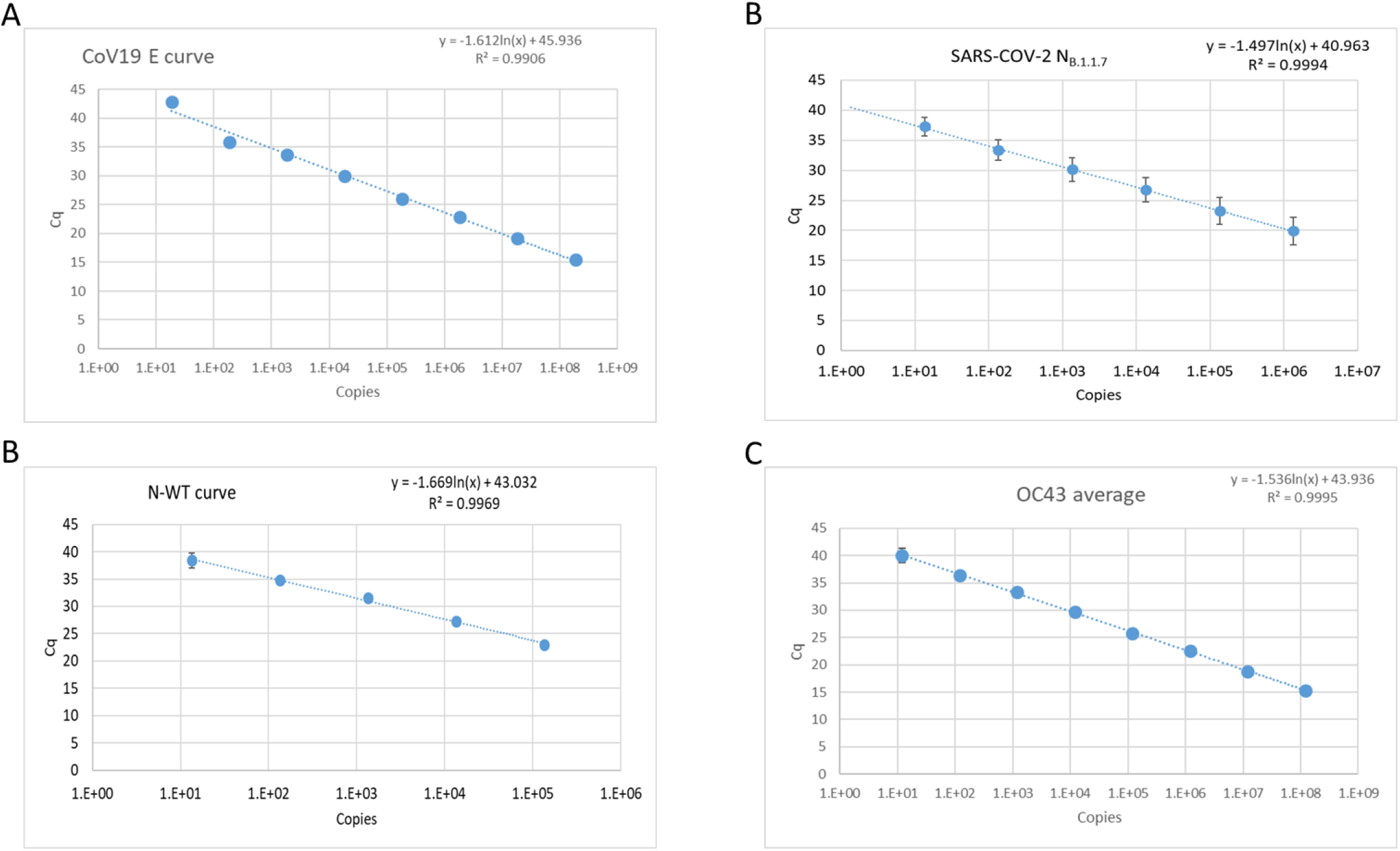
Standard curves of the SC-2 assay reactions. The reaction mix was tested with *In vitro* transcribed RNA molecules containing the assay target sequences. Serial dilutions of each target were tested in triplicates and the resulting regression formula was calculated. (A) E-sarbeco reaction curve. (B) N_B.1.1.7_ reaction curve. (C) N_WT_ reaction curve. (D) CoV OC43 reaction curve. The R^2^ value and the regression formula are shown for each curve.

### 3.3 Analysis of SC-2 variants dynamics using the differential assay

Wastewater monitoring of SC-2 was performed from December 2020 to March 2021 in 11 WWTPs across Israel. WWTPs population catchment area is estimated at about 5.2 million people, which is over 55% of Israel’s population (Supplementary **Table S1**). The WWTPs were clustered into four regions: north, central, Jerusalem, and south. In each region, 3-4 WWTPs were sampled periodically at least once a month. Analysis of the SC-2 dynamics in the northern region showed a mixed trend, where a slight decrease of the total normalized viral load (NVL) was observed in the El Hamra and Safed WWTPs, and a sharp decrease was evident in the Haifa WWTP. Turnover of the variant dominance from WT to B.1.1.7 was completed by the end of February 2021 (**Figure S2A**). Monitoring of the four WWTPs of the central region showed a similar dynamic trend, with total NVL peaking in Jan-21 and slightly decreasing by March. As was in the northern region, by the beginning of March, nearly all samples contained only variant B.1.1.7 (**Figure S2B**). The total NVL values in the Jerusalem region WWTPs showed a similar course, in which total NVL increased until January and then decreased by approximately 90% by March. The variant prevalence switched from 100% WT in December 2020, to 100% B.1.1.7 on the beginning of March 2021 (**Figure S3A)**. Interestingly, while the variant dynamics in the southern region were similar to those in the other three regions, the overall NVL showed a slight increase, from December 2020 until the end of March 2021 (**Figure S3B**). Comparison of the variant prevalence in February 2021 showed that the only WWTP, in which the WT prevalence was above 46%, was El Hamra (99%). In Safed, Ayalon and Rahat, the B.1.1.7 variant consisted between 92% and 99%. The prevalence of B.1.1.7 ranged between 54% and 76% (**Figure S4**).

In order to identify general trends in the variant dynamics, we clustered the results from the WWTPs of each region and examined the dynamics by region, as shown in **Figure 3 and Supplementary Table S2**. The average values obtained for each point were summarized for each region and the percentage of the SC-2 variant was determined (**Figure 3**). The variant turnover was similar in all four regions, switching from 100% WT in December 2020, to 100% variant B.1.1.7 by March 2021. The shift was somewhat delayed in the southern region, where in January, the WT prevalence was almost 100%, compared to 15%-45% in the other regions (**Figure 3**). However, the shift in the southern region was faster compared to the Jerusalem region, 75% and 70% variant B.1.1.7 prevalence, respectively (**Figure 3**). Although the switch from WT to B.1.1.7 was somewhat different in each region, all of them were 100% B.1.1.7 on mid-March (**Figure 3, Supplementary figures S2 and S3**). Comparison of The total NVL showed a moderate but clear decrease in the northern, central and Jerusalem regions, from January to March, while in the southern region, the NVL increased and remained constant during that period (**Figure 3**).

**Figure 3.**
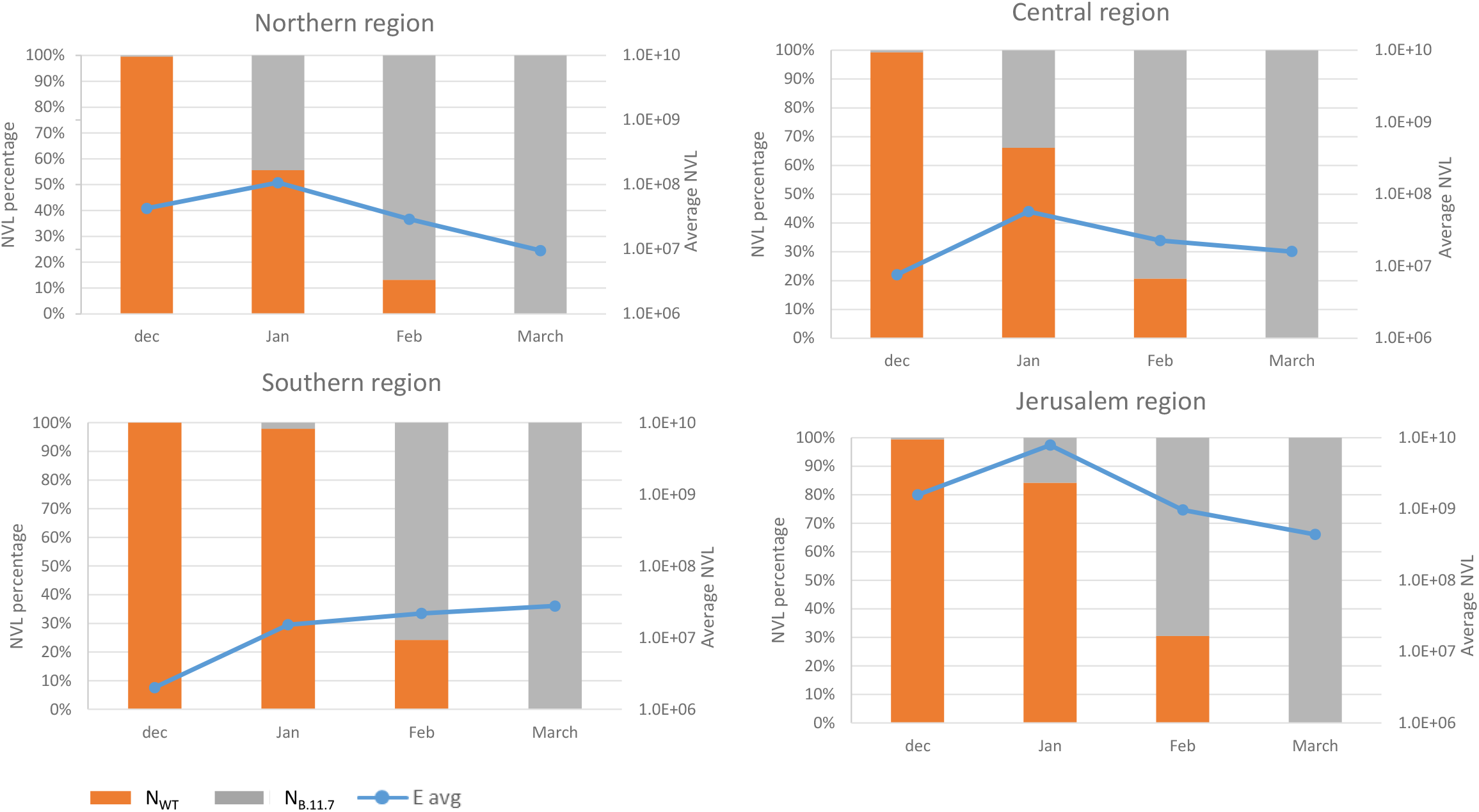
Wastewater SC-2 averaged normalized viral load in different regions of Israel. Blue line represents the averaged amount of E gene NVL in each month in the different regions. The orang and gray bars show the average percentage of Nwt gene and N_B.1.1.7_ gene in every moth in different regions of Israel.

### 3.5 Variant dynamics and total viral load analysis with respect to vaccination status

During December 2020, when the alpha variant was introduced into Israel, a national vaccination campaign was rolled out, using the two-dose BNT162b2 administration (Pfizer). By comparing the wastewater Normalized Viral Load (WW NVL) of each region, we evaluated the possible effect of vaccination on the circulation of SC-2 in wastewater.

The total WW NVL were similar in the northern, central and Jerusalem regions, with a NVL peak on January and a gradual decrease towards March, concurrent with the increase in second dose vaccination rates. This trend was accompanied by a decrease in the new cases reported between January and March (**Figure 4 and Table S2**). The average number of new cases in the northern, central and southern regions was comparable, reaching approximately 1% in January 2021, and decreasing to 0.4-0.6% by March. The average number of new cases in the Jerusalem region was significantly higher, ranging from 3.4% in January, to 0.8% in March (**Table S2**). Although the vaccination rates in the southern region were similar to the other regions, the number of new cases in that region increased, and the total WW NVL elevated accordingly (**Figure 4 and Table S2**). The decrease in total WW NVL and reported new cases in all other regions was parallel to the variant turnover, from WT to B.1.1.7 (**Figures 3 and 4**).

**Figure 4.**
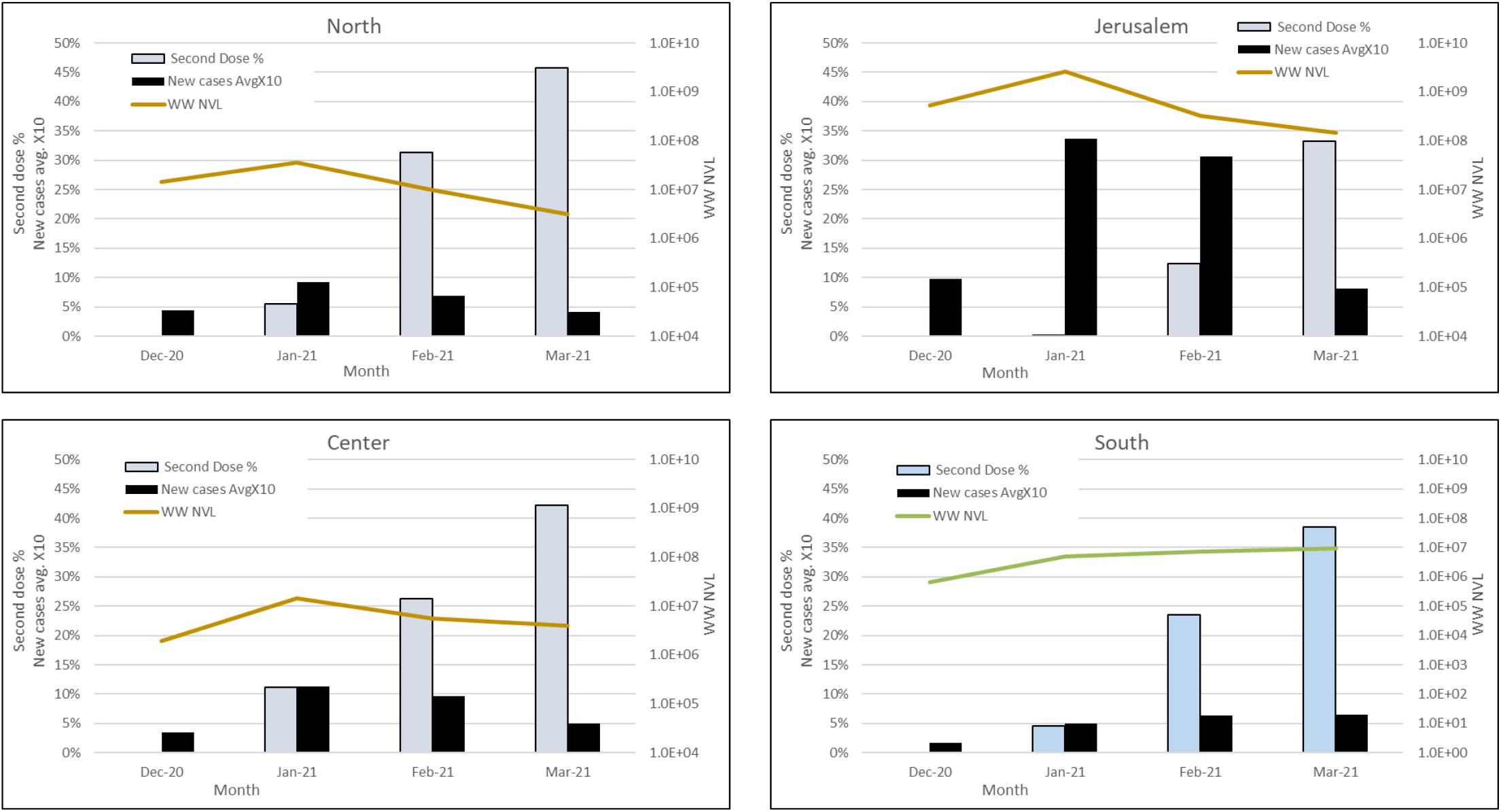
Average of second dose vaccination and new cases rate, compared with wastewater viral load in each region. The 2^nd^ vaccination rates, new SC-2 infection cases and total wastewater normalized viral load (WW NVL) were compared between December 2020 and March 2021, for each region. For clarity of presentation, the percentage of average new cases was multiplied by 10. The WW NVL was calculated as detailed in the Methods section. The vaccination and clinical cases data are from the Israel Ministry of Health website (https://datadashboard.health.gov.il/COVID-19/general).

## 4. DISCUSSION

From the incursion of the SC-2 into Israel, until February 2021, the dominant SC-2 clade in Israel was the Wuhan 19B/20B. The alpha variant was first identified in Israel at the end of December 2020^26^, and by March 2021, it consisted of more than 90% of randomly sequenced clinical samples (Israel Ministry of Health, data not published). Environmental surveillance proved as an important tool in monitoring the COVID19 pandemic in populations and entire large geographic regions^30^. However, information on real-time dynamics of SC-2 variants on a national scale is currently scarce. In this report, we describe the real-time prevalence of circulating SC-2 A19/B19 (Wuhan strain) and B1.1.7 variant, during a period of 4 months, across Israel. Some recent studies of SC-2 variants surveillance relied on the use of genomic sequencing or PCR detection of mutations that are not specific to a particular variant^31,32^. Other studies describe environmental surveillance using a combination of variant-specific and variant-nonspecific reactions^33,34^. Peterson *et al*.^34^ describe the use of six reactions to indicate the presence of alpha, beta (B.1.351) or gamma (P1) variant, but not distinguish between them. The authors examined WW from five major cities, as well as rural communities, determining the prevalence of the three mutations in each site. While the alpha variant was verified by the specific N D3L-directed reaction, as we performed in our study, the other two reactions detected the spike 69-70 and N501Y mutations. These are characteristic, but are not unique, to the alpha variant^35^. Although the authors suggest that identifying these mutations may indicate the presence of beta or gamma variants, there was no definite evidence in the data presented, that this was indeed the case. Furthermore, the need to run a separate assay for each mutation, and the absence of an inclusive control (the “WT” reaction was run for each sample separately, in parallel with the “mutation” reaction) render this approach cumbersome. In our study, we combined an inclusive reaction (E-sarbeco) with the alpha-specific reaction (D3L) and a control reaction (MS-2 phage spiking) to enable rapid and specific screening of a large number of samples. Our comparative analysis shows that the alpha variant emerged in parallel on different sites, in an apparently independent pattern. The first sites, in which alpha variant was dominant by mid-February, were remote from each other (Safed, Ayalon and Rahat, **Figure S4**). This pattern further supports the assumption of independent alpha variant spread. The different variant dynamics during January-February may reflect different infection events at the community level, or simply result from sampling bias, as not all WWTPs were equally accessible for sampling. The complete turnover of all regions on March is consistent with the global variant dynamics, where the alpha variant overtook the WT original SC-2 strain^35^. Analysis of the variant dynamics with respect to population vaccination status showed a correlation between the increase in second dose vaccination rates and the decline in total WW NVL and reported new clinical cases. This occurred regardless of the dominant variant. It is important to note that although the NVL dynamics in the southern region indicated an increase rather than a decrease, the calculated viral load was comparable to that of the northern and central regions, and less than the Jerusalem region. The viral dynamics described in our study are consistent with the recent findings described by Yaniv *et al*.^33^, which showed the turnover of the SC-2 variant during the beginning of 2021, in the city of Be’er Sheva. In a proceeding publication, the authors demonstrated that increased vaccination rates correlate with WW NVL decrease^35^. The authors conducted a thorough monitoring of the variant dynamics in Be’er Sheva and showed that the NVL increased from November 2020 to March 2021, and then started to decrease, following the increase in the second dose vaccination rate^36^. We expanded this examination to 12 different sites across Israel, and found a similar overall change, with local differences between different sites, with evident correlation between the new cases and the NVL values.

In this study, which focused on the SC-2 WT-alpha prevalence, we showed how a real-time and retrospective monitoring of WW could be used to establish the variant dynamics and reflect the population-level response to a national scale vaccination campaign.

Our data show a clear shift in the viral RNA circulation, following the vaccination campaign, thereby demonstrating the importance of wastewater surveillance as an epidemiological tool. Previous reports were limited in the number of sites examined, and mostly used sequencing to identify the circulation of variants of interest^37,38^. A national-scale surveillance in Portugal was comprehensive, but was performed during 2020 and therefore did not examine variant identity^39^. While sequencing provides complete information on the examined sample, it is expensive, not readily amendable for high throughput, and is limited by the sample quality. We recently demonstrated that variant-specific PCR can determine SC-2 variant identity in cases of poor sample quality, where sequencing fails^40^. Here, we show that Variant-specific PCR can be a valuable tool to rapidly identify variant dynamics in WW, in a cost-effective manner, which in turn enables a large-scale screening of WW samples to obtain important epidemiological information. Such approach may be useful in current and future events of variant switch, as is happening now in many countries with the BA.1 variant (Omicron) spreading (Bar-Or and Erster, manuscript in preparation). To our knowledge, this is the first study that describes a national scale SC-2 WW variant surveillance. In addition to serve as an early warning system for the spreading of new variants, it allows a retrospective analysis of variant dynamics and vaccination response, thereby contributing to our understanding of the COVID19 progression and evolution.

## Data Availability

All data produced in the present study are available upon reasonable request to the authors.

## Ethical statement

There was no use of animal or human samples in this study. Therefore, no ethical approval was required. Data regarding population vaccination and new positive SC-2 cases was obtained from the publically available Israel Ministry of Health website at: https://datadashboard.health.gov.il/COVID-19/general

