## Supplemental Material File for "NATIONAL SCALE REAL-TIME SURVEILLANCE OF SARS-COV-2 VARIANTS DYNAMICS BY WASTEWATER MONITORING IN ISRAEL"

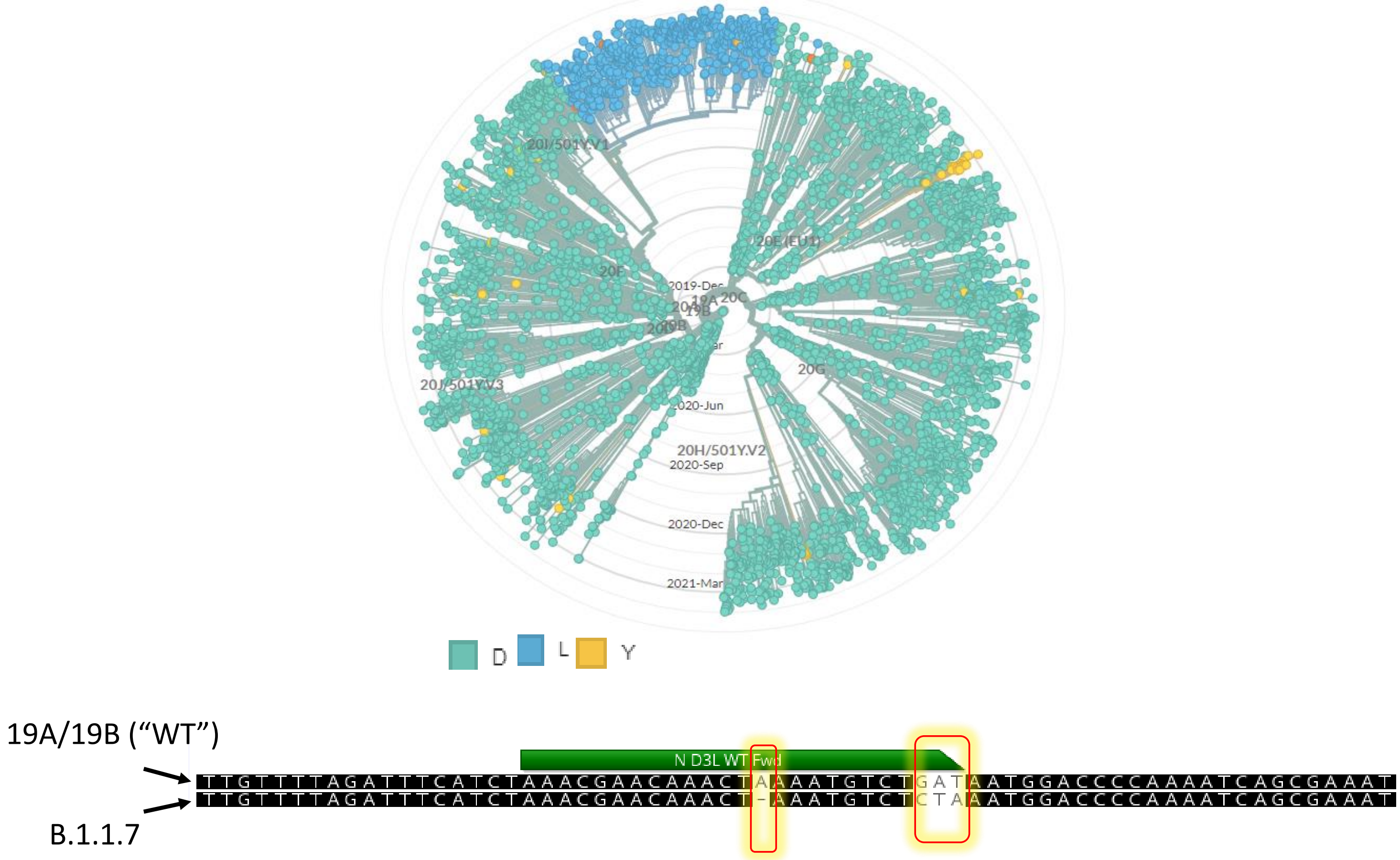


**<u>Figure S1</u>. Design of a differential WT/alpha reaction.**

(A) NextStrain global Analysis of the Nucleocapsid (N gene) sequence showing the uniqueness of the D3L mutation to alpha variant. The dendrogram shows the clades that contains the substitution from D (grey) to L (blue) or Y (orange) in positon 3 of the N protein sequence. (B) Alignment of the specific primer-binding region. The mutations are highlighted.

| WWTP name | Population | Flux (Cubic meters per day) | Region |
|---|---|---|---|
| El Hamra | 21,504 | 2,240 | North |
| Haifa | 583,147 | 100,000 | |
| Zfat | 36,933 | 4,400 | |
| Natania | 291,981 | 39,000 | Center |
| Shafdan | 2,291,901 | 390,000 | |
| Ayalon | 375,649 | 54,000 | |
| Ashdod | 225,939 | 31,100 | |
| Sorek | 873,267 | 108,167 | Jerusalem district |
| Og | 180,000 | 26,586 | |
| Har Homa | 31,250 | 5,000 | |
| Beer Sheva | 272,448 | 39,500 | South |
| Arara | 19,328 | 1,700 | |
| Rahat | 77,335 | 4,500 | |
| **Total** | 5,280,682 | 806,193 | Israel (9,291,000 population) |

**Table S1**. Details of the Wastewater Treatment Plants (WWTPs) sampled during December 2020 to March 2021.

### A. North region

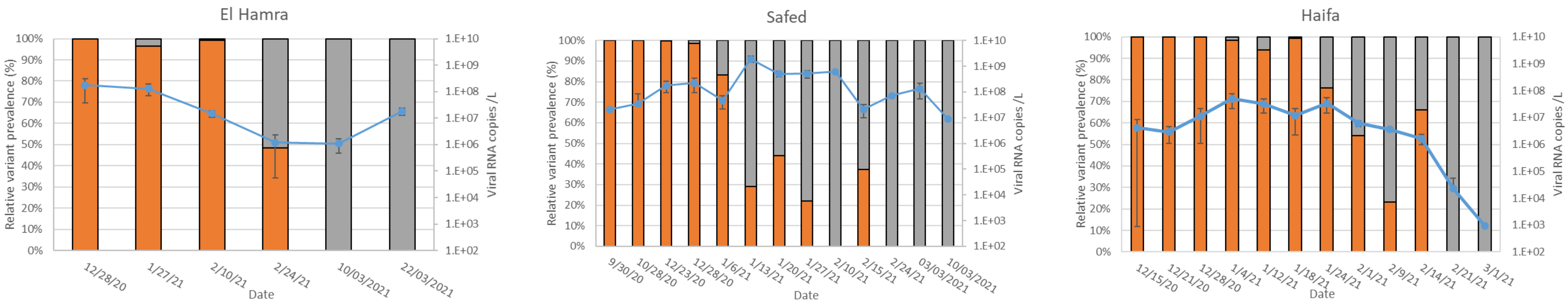


### B. Central region

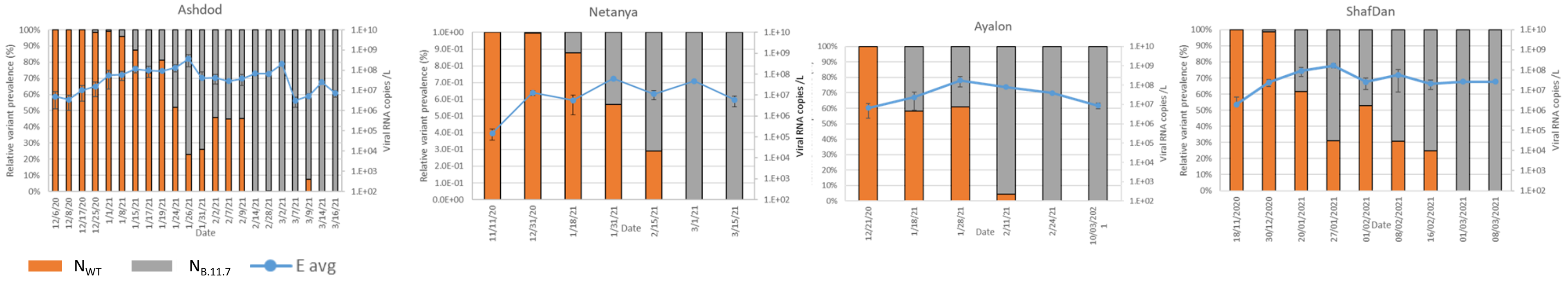


**<u>Fig S2</u>**. SC-2 total load and variant dynamics in the WWTPs of the northern (A) and central (B) regions. The E gene reaction (line with markers) represent the total SC-2 viral RNA copies (left Y-axis). The stacked columns represent the percentages of the WT and B.1.1.7 (right Y-axis).

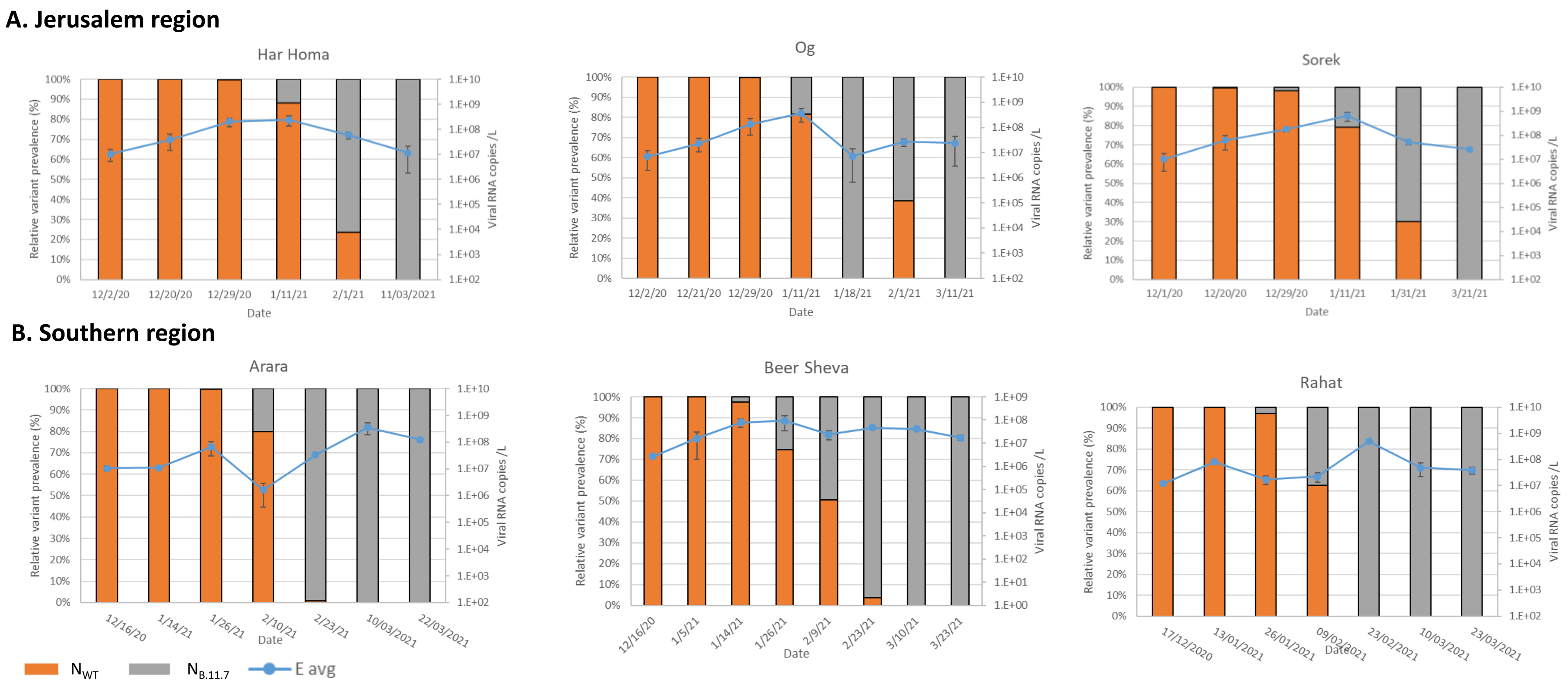


**Fig S3**. SC-2 total load and variant dynamics in the WWTPs of the Jerusalem district (A) and Southern (B) regions. The E gene reaction (line with markers) represent the total SC-2 viral RNA copies (left Y-axis). The stacked columns represent the percentages of the WT and B.1.1.7 (right Y-axis).

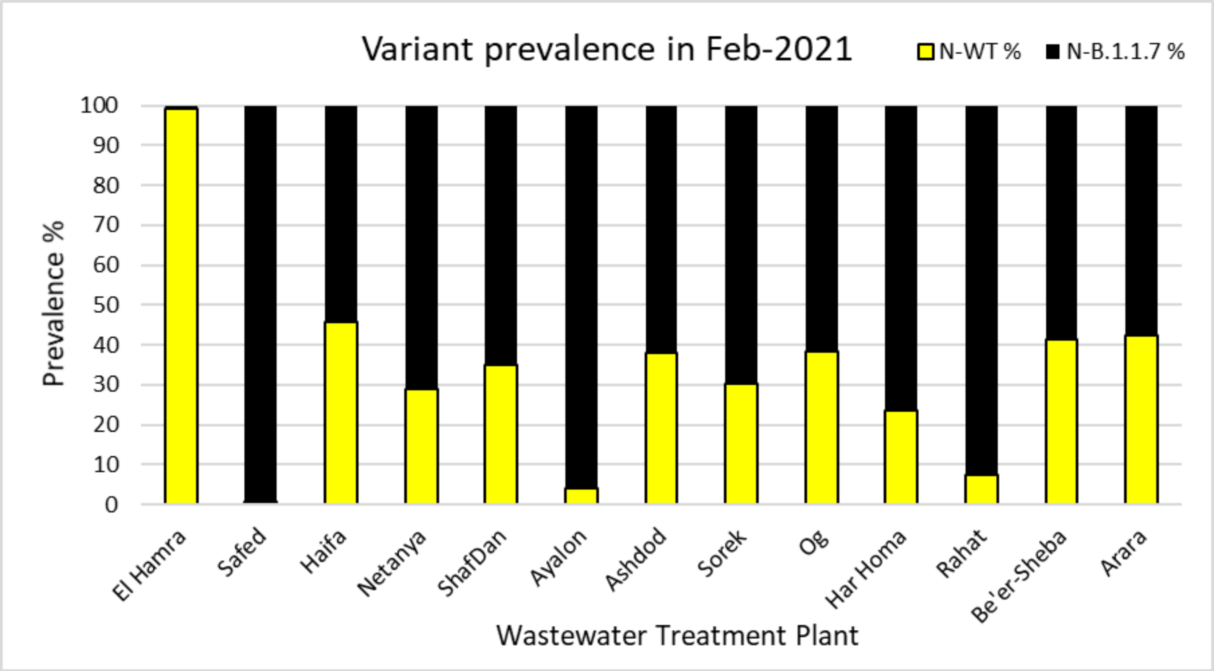


**Fig S4.** Average variant prevalence in each WWTP during February 2021. The average from all the measurements in each WWTP was calculated and the relative percentage of each variant.
